# Crowdsourcing strategies to improve access to HIV pre-exposure prophylaxis (PrEP) in Australia, the Philippines, and China

**DOI:** 10.1101/2023.10.30.23297686

**Authors:** Warittha Tieosapjaroen, Arturo M. Ongkeko, Zhuoheng Yin, Krittaporn Termvanich, Joseph D. Tucker, Weiming Tang, Chunyan Li, Ying Zhang, Nina T. Castillo-Carandang, Jason J. Ong

**Author notes:** **Corresponding author:** Associate Professor Jason J. Ong, Melbourne Sexual Health Centre, Alfred Health, 580 Swanston St, Carlton, VIC, 3053, Australia. Equal first authors. Equal senior authors (Nina T. Castillo-Carandang, Jason J. Ong). **Email addresses of authors:** WT, AMO, ZY, NCC, JJO.

## Abstract

**Introduction:** Many Asian countries have yet to scale up HIV pre-exposure prophylaxis (PrEP). Innovative strategies to increase PrEP uptake are needed. This study examined the use of crowdsourcing to increase PrEP uptake by describing and analysing the experiences of Australia, the Philippines, and China.

**Methods:** Three crowdsourcing open calls were conducted between 2021-2022 in Australia, the Philippines and China. Crowdsourcing has a group of individuals solve all or part of a problem, then share back solutions with the public. All open calls entailed: 1) problem identification; 2) committee formation with local groups; 3) community engagement for idea submission; 4) evaluation of submissions; 5) awarding incentives to finalists; and 6) solution dissemination via web and social media. We examined the number of total and high-quality submissions. We also identified themes across countries.

**Results:** The Australia, Philippines, and China teams received 9, 22 and 19 eligible submissions, respectively. A total of three, 10, and eight submissions had a mean score of 6/10 or greater. Three common solutions emerged across all the finalist ideas: enhancing service delivery; optimizing promotional campaigns; person-centered promotional materials. The winning ideas from the Australian, Filipino and Chinese teams were an anonymous online PrEP service, a printed ready-to-wear garment to create awareness about PrEP, and a poster on PrEP effectiveness, respectively.

**Conclusions:** Crowdsourcing can be a promising and versatile tool for developing PrEP strategies in Asia. Further evaluations via clinical trials can bridge the gap between idea generation and implementation, thus, creating the empirical evidence pivotal for the policy adoption of these innovations.

## Introduction

Approximately six million people were estimated to live with HIV in Asia in 2021, but only 76% of these people were aware of their HIV status. Despite a 21% decrease in new HIV infections in Asia over the past decade, some Asian countries face a dramatic increase in HIV infections.^1^ For example, the Philippines saw a four-fold increase in new HIV cases from2012 to 2021.^2,3^ Similarly, China reported an approximately four-fold increase in new HIV cases from 2010 to 2021.^4,5^ Key populations, including men who have sex with men (MSM), continue to face a disproportionately higher risk compared to the general population.^5^ In contrast, new reported HIV infections were halved from 2012 to 2021. However, the number of new HIV cases has risen among Asian-born men who have sex with men (ABMSM) 54%, from 106 in 2012 to 163 in 2019 before the COVID-19 pandemic.^6^

To control the HIV epidemic, those at risk for HIV must be able to access effective prevention methods, such as pre-exposure prophylaxis (PrEP), an antiretroviral medication-based prevention strategy that reduces the risk of HIV from sex by up to 99% and from injection drug use by at least 74%.^7^ PrEP may be taken daily, “on-demand” (event-driven) or injected every two months, depending on the client’s preference.^8^ However, PrEP uptake in the Australia, China and the Philippines is low.^9^ Strategies to improve PrEP access in each of these countries are warranted since they are at different stages of PrEP implementation and HIV epidemiology (Supplement 1). To do this, looking for new, community-based strategies to facilitate the broader utilisation of PrEP is vital. One such emerging approach is the nationwide crowdsourcing calls for solutions.

Crowdsourcing has a group of people solve all or part of a problem, then share back solutions with the public.^12,13^ This approach has gained traction across numerous disciplines, including public health, where its implementation in HIV programs has demonstrated considerable promise.^14^ Some key advantages of crowdsourcing in public health include expediency, cost-effectiveness,^15^ and its ability to overcome geographic and domain-specific knowledge limitations.^16^ Crowdsourcing could potentially be used to improve HIV prevention, diagnosis, or treatment.^14^ Crowdsourcing has facilitated the development of innovative HIV prevention campaigns, such as a contest in China that generated over 500 novel ideas for HIV prevention, leading to positive outcomes in real-world studies, such as increased HIV testing uptake, enhanced health knowledge, and increased condom use among key populations.^17,18^ These findings highlight the potential of crowdsourcing to enhance HIV programs and improve health outcomes for key populations.

Research gaps remain to be addressed despite the growing evidence supporting the use of crowdsourcing in HIV research. One significant gap is the lack of available information on the logistics and characteristics of crowdsourcing initiatives,^19,20^ which hinders the ability to accurately interpret findings and replicate its success. A more detailed report on the precise mechanics of crowdsourcing efforts would contribute to a better understanding of the factors that drive the success of these initiatives.^20,21^ Additionally, while existing research provides insights into the benefits of crowdsourcing,^18^ it is crucial to describe the local conditions and contexts under which its use is most effective.^21,22^ This paper aims to contribute to the growing literature on the use of crowdsourcing to generate solutions to increase PrEP uptake by describing and analysing the experiences of Australia, the Philippines, and China. This paper offers insight into the various contexts where crowdsourcing can play a significant role in enhancing HIV prevention and control efforts in countries with different stages of PrEP implementation and HIV epidemiology.

## Methods

The HOPE network organised the open calls conducted in Australia and the Philippines (HOPE-endHIV.com). This network connects regional experts in Australia, the Philippines, and Thailand to reduce HIV transmission by optimising PrEP in East Asia, including generating new knowledge and solutions to optimise the use of PrEP, and translating research and community insights into accessible forms for PrEP program and policy development. The open call in China was conducted by Social Entrepreneur to Spur Health (SESH), an organisational partner of the HOPE network. All open calls in Australia, China, and the Philippines were conducted based on the World Health Organisation’s Crowdsourcing in Health practical guide^12^ as follows: 1) identify problems; 2) set up steering and/or organising committees; 3) engage the local community to submit their ideas to open calls; 4) receive and evaluate submissions; 5) recognise finalists; and 6) share and implement solutions.^23^ The steering committee supervised the process and ensured adherence to the protocol, while the organising committee handled the daily crowdsourcing operations, from promotion to preparing entries for judging. All judges received a full description of the judging criteria in advance and were asked to provide a single score for each submission on a 10-point scale, where higher scores refer to better quality. All open calls were tailored to each country’s unique HIV epidemiology and culture. A summary of each country’s open call process is presented in Figure 1. Below, we discuss in depth how each country conducted its crowdsourcing activities.

**Figure 1.**
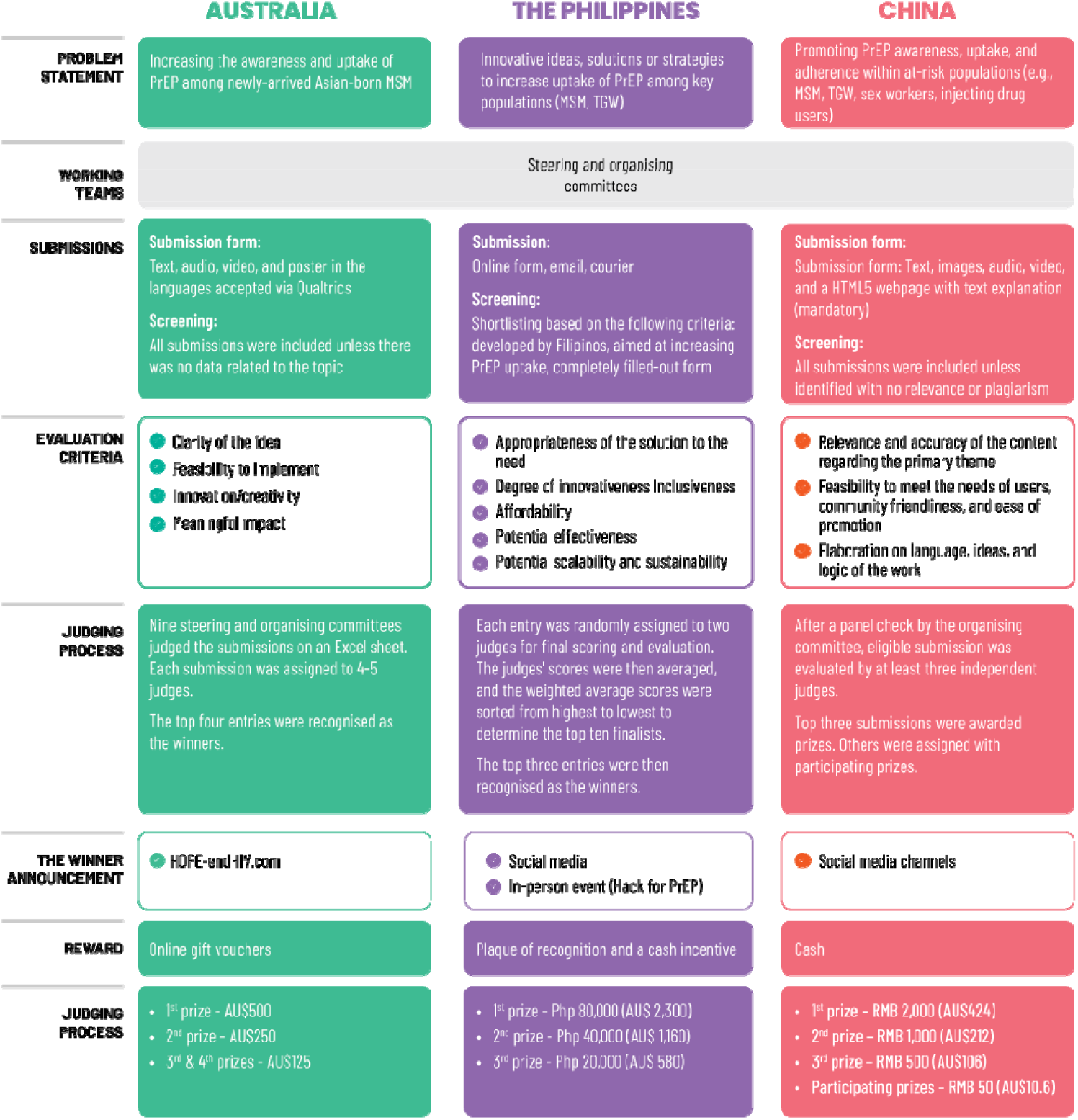
Summary of the Open Call Process in Australia, the Philippines, and China.

### Australia

Our Australian open call aimed to increase the awareness and uptake of PrEP among newly arrived Asian-born men who have sex with men (MSM), conducted between March and October 2022. The steering and organising committees comprised clinicians, researchers, end users, policymakers, and members of local organisations. The open call was advertised online via social media platforms (such as Facebook) and local organisation email lists, group chats, websites, and Facebook fan pages between August and September 2022 in six languages (i.e., English, Thai, Chinese, Tagalog, Bahasa Indonesian and Vietnamese).^24^ We used a survey platform, Qualtrics (Qualtrics, Provo, UT), for receiving submissions. The total advertising cost was AU$ 7,465. Each individual could submit multiple ideas. The social media plan can be found in Supplementary 2. We accepted a 500-word message, an A-1 poster, a 3-minute video, or audio recordings by an individual or a team in any of these six languages which participants were familiar with (i.e., English, Chinese, Thai, Bahasa Indonesia, Hindi, Punjabi, Vietnamese, or Filipino/Tagalog). There were no restrictions on the gender, nationality, and age of participants. Nine steering and organising committee members judged the submissions on an Excel sheet. Each submission was assigned to 4-5 judges. The judging criteria included: 1) clarity of the idea, 2) feasibility to implement, 3) innovation/creativity, and 4) meaningful impact. The finalists received AU$500 (1st prize), AU$250 (2nd prize), and AU$125 (3rd and 4th prize), and the result was announced via the open call website.^24^

### The Philippines

Our Philippines open call aimed to increase PrEP use among key populations, including MSM and trans women (TGW), conducted from October 5 to November 2, 2022. The project team formed two committees: the organising and steering committees. The organising committee comprised of eight project team members. The steering committee consisted of six members, including clinicians, academics, and representatives from the LGBTQIA community. The project team used a dedicated social media page to promote the call.^25^ informed by a comprehensive social media plan (Supplement No. 3) and by forming partnerships with key government agencies. We accepted submissions through email, online forms, and direct office submissions to ensure wide accessibility. Entries underwent two screening stages: an initial eligibility check by the organising committee and a detailed evaluation by an expert panel, including the steering committee and three additional experts. During the eligibility screening, the organising team reviewed the entries to see if they were developed by Filipinos, focused on increasing the uptake of PrEP, and most importantly, whether the submission forms were completely and accurately filled out to provide sufficient information for a fair review. Criteria for final evaluation included appropriateness, innovation, inclusivity, affordability, and potential effectiveness. Two judges from the expanded steering committee scored each entry, with the top ten entries shortlisted. The top three innovations received plaques and cash prizes of Php 80,000 (AU$ 2,300), Php 40,000 (AU$ 1,160), and Php 20,000 (AU$ 580), respectively. Winners were notified via email about their awards. The finalists were featured on the HOPE Philippines’ social media page.^25^

### China

The China crowdsourcing open call focused on soliciting submissions to enhance PrEP awareness, uptake, and adherence within at-risk populations. This initiative spanned from March to May 2021. The steering and organising committees and judging panel consisted of community-based organisation (CBO) members, healthcare professionals, and academics, who were selected based on their expertise in sexual health and crowdsourcing. Eligible submissions included text (concepts, slogans, short articles, < 1000 characters), images (photographs, posters, logos, drawings), and multimedia content (videos, audio, HTML5 webpage, < 3 minutes) required with an explanation of fewer than 300 words. All announcements related to the open call (promotions, participating instructions, deadlines, prizes) were channelled through the social media platforms of the holders and CBOs in different provinces. The online survey we used for receiving submissions was included in Supplementary 4. During the submission period, online assistance via email or chat messages was open to all participants for inquiries. All submission entries were screened to check for relevance to our open call and to identify plagiarism. Then, each product was evaluated by at least three independent judges. The quality of the entries was judged on 1) the relevancy of the idea, 2) feasibility to implement, and 3) aesthetic design and elaboration of details. The award structure consists of a single first, second, and third prize, each accompanied by a monetary incentive of 2000 RMB (AUD$ 424), 1000 RMB (AUD$ 212), and 500 RMB (AUD$ 106) respectively. Besides, to encourage active engagement, an AUD$ 10.6 incentive were set additionally as an award to 20 participants. The final list of winners was announced through public media channels.

Ethics approval for this study was obtained from the Alfred Hospital Ethics Committee, Melbourne, Australia (project number 266/22). All participants were provided with a participant information sheet and consent implied by the submission of ideas.

## Results

All countries received eligible submissions, successfully evaluated submissions and obtained finalists. Three themes emerged across all the finalist ideas from Australia, the Philippines and China as follows: 1) service delivery, including an anonymous online PrEP service, community-based mobilisation algorithm, and PharmAssist for PrEP; 2) promotional campaigns, including project SHAFT (Sustainable HIV & AIDS Awareness through Fashion Tales) and social media campaign for Vietnamese MSM living in Australia; and 3) promotional materials, including posters on PrEP use, effectiveness, and comics on PrEP and other STIs (Figure 2).

**Figure 2.**
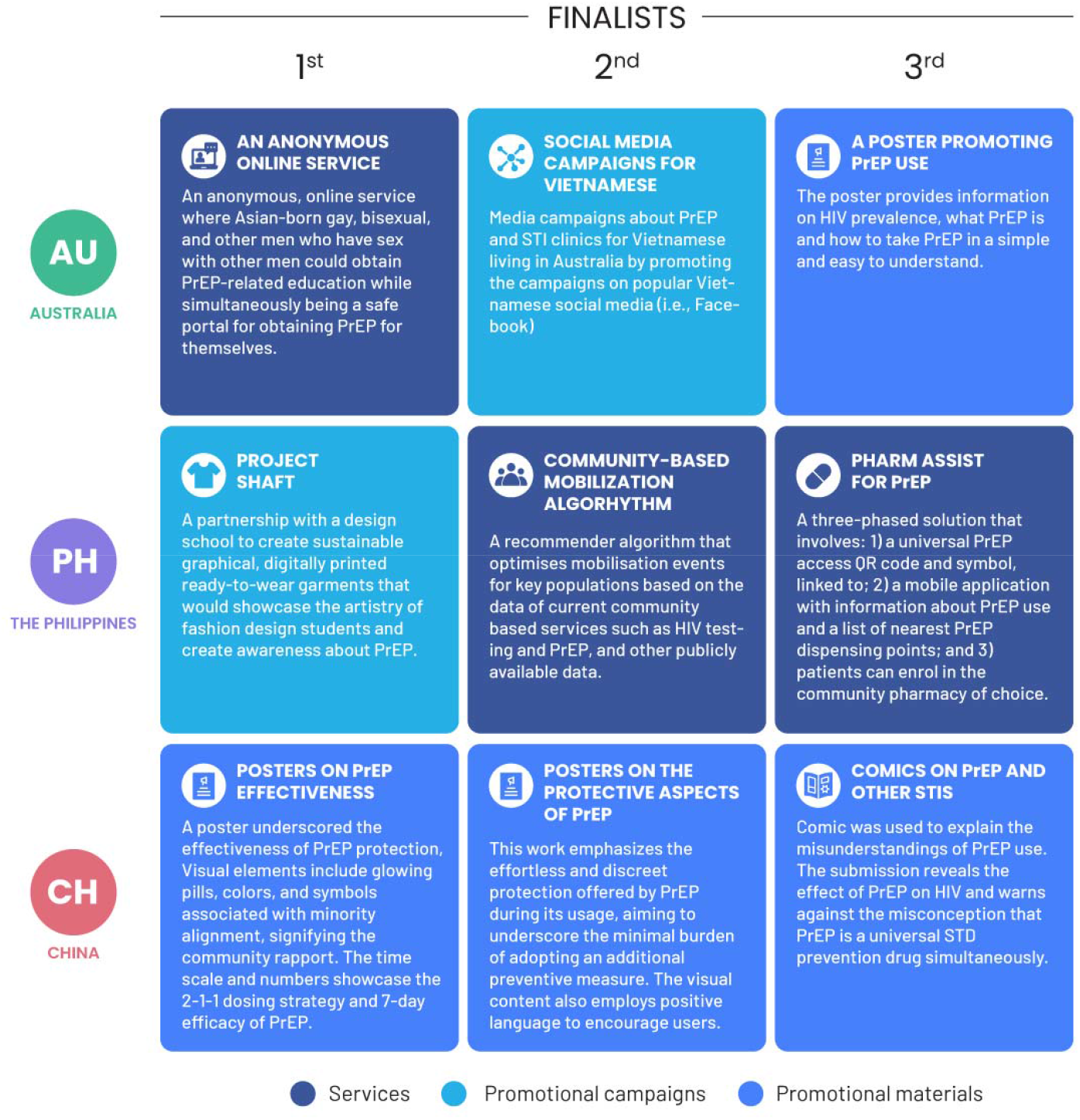
The Summary of the Top Three Finalist Ideas from Australia, the Philippines and China.

### Australia

The advertisement via a social media platform (i.e., Facebook) reached 312,823 users, and 3,962 clicked on the open call post within 2 months of the call launch. We received 11 submissions, with nine submissions eligible for judging (Figure 3). The submissions included text and infographics. Two submissions were excluded due to no relevant data. All submissions were in English. There were nine participants: three males, two females, and four cisgenders, with a mean age of 34. Six (67%) participants were born overseas. The median score for all submissions was 22.8/40 (IQR=17.8, 24.8, range=10.8,26.8). The score sheets are shown in Supplementary 2. The summary of the top three finalist ideas is shown in Figure 2, and the full submissions can be found in Supplementary 2. After the open call, all four finalist ideas were further developed during our face-to-face designathon in April 2023.^26^ The winning prototype from the designathon is undergoing testing in an ongoing clinical trial.

**Figure 3.**
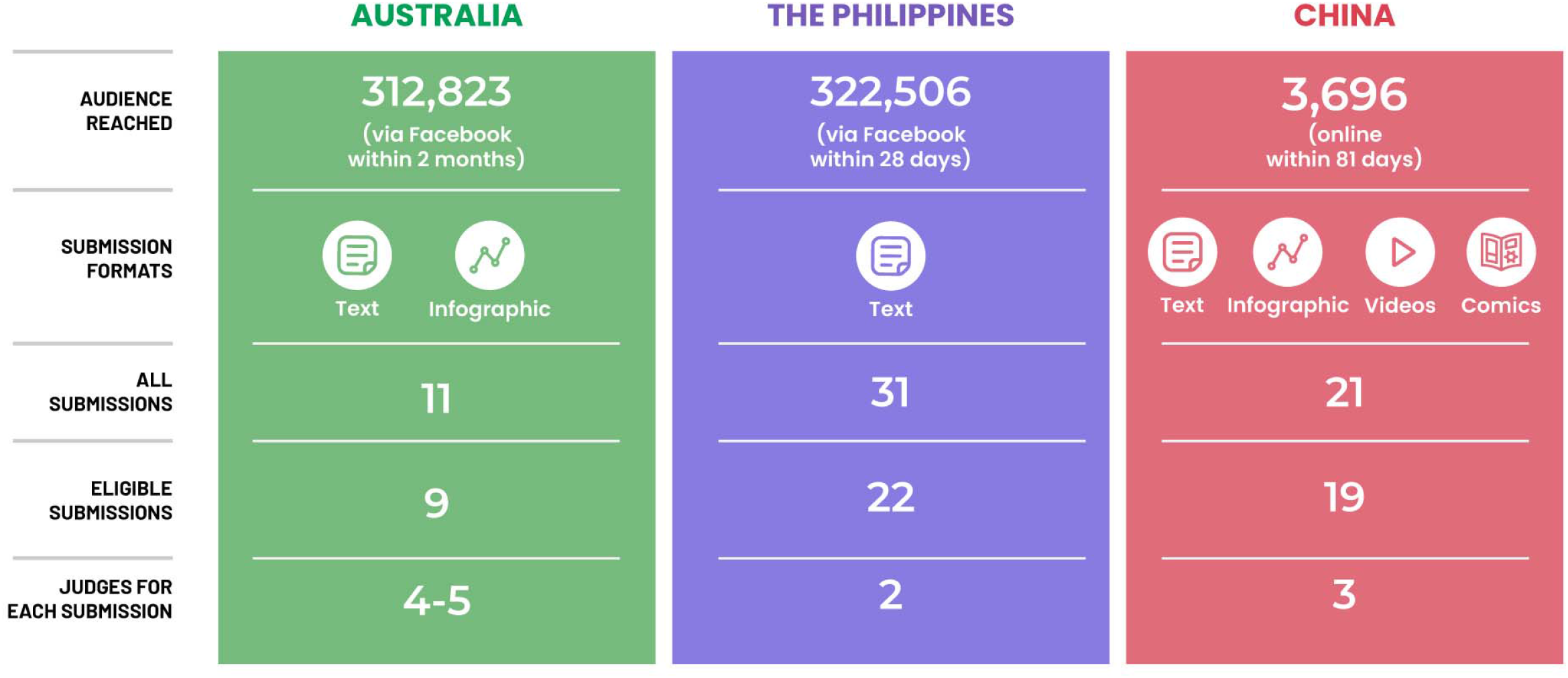
The open call outputs from Australia, the Philippines and China teams.

### The Philippines

The project reached 322,506 people and generated 8,523 post engagements through Facebook within 28 days of the call launch. The call attracted 31 submissions, with initial screening resulting in 22 eligible entries (Figure 3). The nine excluded submissions lacked sufficient information or were deemed irrelevant to the call’s objectives. Eligible entries were randomly assigned to two judges for final evaluation, and the average of their scores was used to rank the entries. The median score for all submissions was 3.8/5 (IQR=3.6, 4.0, range=3.5, 4.4). The score sheets are shown in Supplementary 3. A brief description of the top three innovations is presented in Figure 2, whereas the rest of the finalists can be found in Supplementary 3.

### China

A total of four official online promotions were published in 81 days with 3,696 audience reached. Nineteen submissions were eligible of total of twenty-one received. All independently evaluated by the judging panel (Figure 3). Eight submissions were in text or slogans, two were videos, and the remaining nine were posters, photos, or comics. The judging panel selected five posters and one comic as the most distinguished contributions, with three also securing participating prizes. The median score for all submissions was 29.2/50 (IQR=22.9, 34.1, range=6.0, 40.2). The score sheets are shown in Supplementary 4. The top three innovations are described in Figure 2, and original submissions are included in Supplementary 4. After the open call, the six revised output were delivered to Chinese PrEP users in a PrEP demonstration trial biweekly to test its preliminary effects on adherence enhancement further.^27^

## Discussion

This paper reported the experiences and insights from implementing crowdsourcing calls to promote PrEP utilisation in Australia, the Philippines, and China. This study demonstrates the potential of crowdsourcing as a tool applicable across different countries, cultures, health systems, and HIV epidemics. Using crowdsourcing, we engaged communities to create and develop locally-informed solutions. This paper contributes to the existing literature by showcasing real-world examples of crowdsourcing applications in developing strategies to end HIV transmission. We also discuss the challenges we encountered during our open call and how to best address them\.

Crowdsourcing approaches helped engage key populations in three diverse settings and develop messages for locally tailored PrEP campaigns. In all three countries, crowdsourcing meaningfully engaged communities to address persistent challenges (e.g., stigma and discrimination) encountered by vulnerable populations, such as people living with HIV and MSM.^28-31^ By enabling collaborations with people from diverse backgrounds, experiences and skills and by encouraging community engagement, crowdsourcing allows for out-of-box thinking and increases community ownership.^23^ All these solutions highlight the potential of crowdsourcing to solicit solutions to improve awareness, acceptance, and access to PrEP in each country contributing to efforts to end HIV transmission. Given the promising outcomes, the next logical step is determining how to scale up these innovations effectively.^33^ The scale-up of these innovations can be facilitated in collaborative environments such as designathons.^34^ These events cultivate creativity and teamwork and offer a framework to operationalise and implement these solutions in real-world contexts. As we bridge the gap between idea generation and implementation, clinical trials alongside economic evaluations are important to produce the empirical evidence pivotal for the policy adoption of these innovations.^35^ Two ongoing clinical trials in China utilise community-derived interventions from crowdsourcing to achieve similar goals.^27,36^ The Australian team is in the process of implementing their solution in a clinical trial.

Parallel to these more traditional next steps, the convergence of arts and sciences and popular culture emerges as a promising avenue to test and upscale PrEP, especially in the popular culture of countries in Southeast Asia like the Philippines and Thailand. Beauty contests, deeply rooted in the social fabric of these countries, serve as more than just showcases of physical beauty. They are seen (among others) as platforms for social mobility, personal expression and development, and national recognition.^37^ More significantly, these contests provide the often marginalised transgender community with platforms to express their identity, gain acceptance, and advocate for their rights. In the Philippines, these contests feature prominently in town fiestas (conceived initially as religious public festivals with various processions and activities to honour a patron saint) and public events, drawing large audiences and fostering broader acceptance of the transgender community. In Thailand (one of the 3 HOPE Network countries which is currently planning for its own crowdsourcing call), the high-profile, nationally broadcast transgender beauty contests offer a powerful stage to engage key populations. Given these events’ widespread reach and high engagement, especially among key populations, they present a potentially useful platform for promoting PrEP. The inclusivity of these contests could foster PrEP awareness and acceptance. As a form of personal expression, fashion can be harnessed to raise awareness, combat stigma, and advocate for the acceptance of PrEP. Exploring these intersecting elements presents a new research area that can enhance our understanding of health equity within a broader cultural context, encouraging a more inclusive discourse on health promotion.^38^

Challenges observed during crowdsourcing implementation included recruitment, social media management, quality of submissions, and the judging system. In the Philippines, recruitment via social media specially for a short period of time (less than one month) was not successful in reaching diverse participants and may have contributed to several submissions being similar. Our data are consistent with research from TDR^12^ suggesting that promotion of diverse participants is one key driver of success for crowdsourcing activities. Meanwhile, social media management encountered some difficulties in Australia. Despite advertisements targeting Asian-born MSM in Australia, a multicultural country, there were people reacted on the Facebook ads in a discriminatory way, which is a common risk for any online public campaign. Consequently, proactive measures were taken to maintain a positive campaign atmosphere. Additionally, Australia saw some level of bias from the judges given that they came from diverse backgrounds with different preferences, and such bias was mitigated by assigning at least three judges for each submission. Lastly, some submitted entries were excluded due to irrelevancy. This could be caused by inadequate and unclear instructions to participants from the start. Clearer communications, using guiding questions to frame details of entry, could benefit the implementation.

The main strength of our paper is to present details of how crowdsourcing was implemented in three different settings but with the same goal of improving access to PrEP. We showcase the success and challenges of implementation in these settings. Our study should be read in light of some limitations. First, our project originally expected a crowdsourcing open call from our Thailand team, but they have yet to implement their crowdsourcing activities due to challenges in securing funding. The implementation plans for Thailand can be found in Supplementary 5. Second, there could be selection bias in excluding individuals who may not have access to or were not active on platforms used for recruitment. This may limit the diversity and representativeness of the submissions. Third, although efforts were made to advertise the open call in non-English languages in Australia, we did not receive any non-English submissions. This language bias could result in the underrepresentation of certain communities or individuals who are not proficient in the advertised languages. Finally, our findings are specific to the contexts of Australia, the Philippines, and China, but some of the principles shared may still apply to other contexts or populations with similar cultural, social, or healthcare contexts.

## Conclusions

In conclusion, crowdsourcing is a promising and versatile tool for generating community-based solutions. Involving a diverse range of participants, including those as part of the populations to reach, enabled the generation of innovative ideas and solutions to increase the awareness and uptake of PrEP among specific populations. Using online platforms and strategic partnerships with key stakeholders was crucial in promoting the open call. This indicates the value of leveraging digital platforms and community networks to increase engagement and participation.

## Data Availability

The data supporting this study's findings are available from the corresponding author upon reasonable request.

## Authors’ Contributions

JJO, JT, NP and WTang conceived the ideas. WTi, AO, KT, ZY, NTC and JO wrote the first draft of the manuscript. WTi revised and finalised the manuscript. All authors contributed to the manuscript and approved the final version for submission.

## Acknowledgements

We thank ACON, Australian and New Zealand Tongzhi Rainbow Alliance, Study Melbourne, Thorne Harbour Health, Positive Life NSW, National Association of People with HIV Australia, International Student Health Hub, Ethnic+ in Queensland, Australian GLBTQ Multicultural Council, Forcibly Displaced People Network, International Student Sexual Health Network, Selamat Datang, Trikone Queer South Asians in Melbourne, Switchboard Victoria and Victorian Pride Centre, University of New South Wales Health Promotion, Limin Mao, and Judith Dean for their assistance in promoting the open call in Australia. In the Philippines, our gratitude extends to our three pilot implementation sites: LoveYourself, Inc. (Mandaluyong City), The Rajah Community Center by the Family Planning Organization of the Philippines (Iloilo City Chapter), and the Save and Improve Lives (SAIL) Clinic in Calamba City facilitated by the Sustained Health Initiatives of the Philippines (SHIP, Inc). The Philippine team further wishes to acknowledge the Republic of the Philippines’ Department of Health, and the Philippine Council for Health Research and Development (PCHRD) of the Department of Science and Technology; and the University of the Philippines Manila for their support in promoting the call. Special thanks also go to the Communications Team of the HOPE Philippines, particularly Ms. Jean Francis A. Barcena and Ms. Korinna C. Looi, RN for their instrumental work in developing the strategic communications plan and materials.

