## Supplementary for "Crowdsourcing strategies to improve access to HIV pre-exposure prophylaxis (PrEP) in Australia, the Philippines, and China"

**Supplementary 1. HIV epidemiology in Australia, the Philippines, and China**

In Australia, it was estimated that 29,640 were living with HIV in 2021, with less than 100 AIDS-related deaths.^1^ The Therapeutic Goods Administration approved pre-exposure prophylaxis (PrEP) for HIV prevention indication in 2016.^6^ After a study demonstrated a reduction in HIV transmission among who have sex with men (MSM) as part of a PrEP implementation trial in New South Wales,^7^ PrEP was subsequently listed on the pharmaceutical benefits scheme (PBS) in 2018 for Medicare-eligible individuals identified as at risk of HIV infection (e.g., condomless intercourse with casual male partners, methamphetamine use and an STI diagnosis).^8^ Medicare is the publicly-funded universal health care insurance scheme in Australia. Five years later, Australia had the highest number of PrEP access per capita globally, with more than 62,000 accessing PrEP.^9^ Australia reported a 52% decline in new HIV infections from 1,068 in 2012 to 552 cases in 2021. However, the number of new HIV cases has risen among Asian-born men who have sex with men (ABMSM) 54% (From 106 in 2012 to 163 in 2019).^10^ A national Gay Asian Men survey in Australia reported a lower PrEP use among ABMSM, especially those from South-East (aOR=0.47, 95%CI 0.29-0.74, P=0.001) and South Asia (aOR=0.36, 95%CI 0.18-0.82, P=0.004), compared to gay Asian men born in Australia (Unpublished data). ABMSM reported additional barriers to PrEP, including insufficient knowledge about PrEP and HIV back in their country of origin, low-risk perception of HIV, and no subsidized PrEP from the government.^11,12^ Medicare-eligible individuals can purchase a 30-day PBS PrEP pack starting from AU$28. Meanwhile, the total cost of accessing PrEP for Medicare-ineligible individuals can be much higher as they have to pay out-of-pocket costs for general practitioner consultations and laboratory tests every three months (~AU$200). Individuals without Medicare have to either purchase PrEP at full price (AU$15-65 per month) or import cheaper PrEP from overseas pharmacies.^13^

The prevalence of HIV in the Philippines remains a significant public health concern, as the country is facing one of the fastest-growing HIV epidemics globally.^3^ According to recent data from the Department of Health (DOH) HIV/AIDS & ART Registry of the Philippines (HARP), approximately 116,504 people were living with HIV as of May 2023, with 1,256 newly diagnosed HIV cases (26% of whom had an advanced HIV infection at the time of diagnosis) reported in May alone.^14^ The report further highlights that most new HIV diagnoses were among males, accounting for 94% of all reported cases, with the most affected age group being 25-34 years old, constituting 46% of reported cases. The most common mode of HIV transmission was sexual contact, with male-to-male sexual contact accounting for 70% of all reported cases. Moreover, the report underscores a continued increase in the number of AIDS-related deaths, which has reached a total of 6,709 since 1984. The integration of PrEP into the healthcare services of the Philippines began six years ago. This initiative took off in 2017 when LoveYourself (<https://loveyourself.ph/>), in collaboration with the Department of Health (DOH), launched a two-year study. The objective was to assess the community-based delivery of PrEP for men who have sex with men (MSM) and transgender women. In this cohort of 240 participants, no new cases of HIV infection or side effects were recorded. These encouraging findings propelled the DOH to expand the use of PrEP across the country.^15^ In 2021, the DOH issued temporary guidelines for implementing PrEP. A year later, the Health Technology Assessment Council (HTAC) endorsed the inclusion of oral PrEP in the Philippine National Formulary.^16^ These guidelines highlighted the critical need for PrEP awareness campaigns. They also urged the utilization of HIV healthcare provider networks to promote PrEP. They emphasized facilitating access to PrEP, improving the provision of PrEP, and refining client referral systems for PrEP. Oral PrEP costs around 1,500 pesos (approximately AU$40) per bottle, equating to 50 pesos per tablet.^17^ Its availability is mainly limited to select regions of the country, particularly Metro Manila.^18^ However, according to the DOH guideline, PrEP medications can be supplemented using resources from HIV projects and grants. Additionally, local government units are authorized to procure PrEP drugs based on their yearly operational plans.

China recorded 6,0154 new HIV infections in 2021 alone, reaching a total of 1,053,000 people living with HIV.^19^ Key populations, including MSM, continue to face a disproportionate higher risk compared to the general population.^19^ In 2019, China issued the Implementation Plan for Curbing HIV Transmission (2019-2022),^20^ emphasizing the need to pilot test PrEP, particularly among key populations. Despite the availability of low-cost generic PrEP in the market and a high reported willingness for PrEP,^21,22^ the actual uptake rate remained extremely low in China. A cross-sectional survey conducted among 1915 MSM from 34 Chinese in 2023 reported only 23 (1.2%) ever had PrEP-using experiences.^23^ Moreover, lower adherence intention was observed among MSM in China.^21^ Previous studies have identified both structural constraints and individual-level barriers contributing to the predicament, including lack of effective PrEP messaging, no coverage of insurance, affordability, high gay- and HIV-related stigma in clinic settings, shortage of PrEP providers, and individual’s concerns over side effects, preference for condoms, and lack of engagement with HIV prevention care overall.^24^

**Table S1. The summary of current HIV and PrEP situations in Australia, the Philippines, and China**

|  | **Australia** | **Philippines** | **China** | **Thailand** |
| --- | --- | --- | --- | --- |
| **Estimated number of new HIV cases reported** | 552 cases per year in 2021^10^ | 21,000 cases per year in 2021^1^ | 60,154 cases per year in 2021^19^ | 6,500 cases per year in 2021^1^ |
| **Number of people newly diagnosed with HIV** (selected years) | 1,068 cases in 2012 to 552 cases in 2021 (52% reduction)^10^ | 5,000 cases in 2010 to 21,000 cases in 2021 (420% increase)^3^ | 15,982 cases in 2010 to 60,154 cases in 2021 (376% increase) ^19,36^ | 16,000 cases in 2010  11,000 in 2015  6,500 cases in 2021^1^ |
| **Total number of people living with HIV** | 29,640 are living with HIV in 2021^10^ | 140,000 people living with HIV in 2021^3^ | 1,053,000 people living with HIV in 2021^37^ | 520,000 people living with HIV of all ages in 2021^1^ |
| **Total number of HIV/AIDS deaths** | <100 deaths in 2021 ^1^ | 1,200 deaths in 2021^1^ | 19,623 deaths in 2021^19^ | <100 deaths in 2021^1^ |
| **Target number of clients, and number of people receiving PrEP** | PrEP users: 39,643 (December 2022)^38^  Target PrEP users: unspecified | PrEP users: 12,906 (December 2022)  Target for 2026: 150,000 | PrEP users: Less than 82,260 (in 2019)^39^ | PrEP users: 58,269 (June 2023)  Target for 2023: 147,534^32^ |
| **Coverage by the national health insurance program** | Covered for Medicare-eligible individuals | Not covered by the Philippine Health Insurance Corporation (PhilHealth) | Not covered by the Chinese Health Insurance Corporation | Covered for all Thais |

PrEP=Pre-exposure prophylaxis

**Supplementary 2. Additional details on the open call in Australia**

**Table S2. The purposed social media advertisement plan for the open call in Australia**

|  | **Sent the open call materials to all the organisations/followed up** |  |  |  |  |  |  |
| --- | --- | --- | --- | --- | --- | --- | --- |
| **July** | **25** | **26** | **27** | **28** | **29** | **30** | **31** |
|  | 1. FB post graphics 2. Open call advertisement for websites 3. FB Ads instruction 4. Website link |  |  |  |  |  |  |
|  | **Tested out the campaign setting 1 on FB and Instagram** |  | **The organisations shared the ads**  **Start the FB Ad campaign- set a budget - $500 daily for running the ad for four days** | **Checked responses/clicks** |  |  |  |
| **August** | **1** | **2** | **3** | **4** | **5** | **6** | **7** |
|  | 1. FB post graphic 2. FB story graphic |  |  | Looked at the demographic characteristic of people who click on the post and narrow the target audience down |  |  |  |
|  | **Checked responses/clicks** |  |  | **Checked responses/clicks** |  |  |  |
| **August** | **8** | **9** | **10** | **11** | **12** | **13** | **14** |
|  | Adjust the target audience |  |  | We received less than 50 responses-discussed about FB live Q&A session |  |  |  |
|  |  | **The organisations reposted on FB (due date in 2 weeks)** |  |  | **Stopped FB Ads and discussed with the team** |  |  |
| **August** | **15** | **16** | **17** | **18** | **19** | **20** | **21** |
| **August** | **22** | **23** | **24** | **25** | **26** | **27** | **28** |
|  |  |  |  | We received less than 50 responses-extend Facebook Ads |  |  |  |
| **August** | **29** | **30** | **31** | **September 1** | **2** | **3** | **4** |
|  |  | **The organisations reposted on FB (we moved deadline)** |  |  |  |  |  |
| **September** | **5** | **6** | **7** | 8 | 9 | **10** | **11** |
|  |  | **Checked responses/clicks** |  |  |  |  |  |
| **September** | **12** | **13** | **14** | 15 | 16 | **17** | **18** |
|  |  | Adjust the target audience |  |  |  |  |  |
|  |  | **Checked responses/clicks** |  | **The organisations reposted on FB (we moved deadline)** |  |  |  |
| **September** | **19** | **20** | **21** | 22 | 23 | **24** | **25** |
|  |  | Adjust the target audience |  |  |  |  |  |
| **September** | **26** | **27** | **28** | 29 | 30 |  |  |

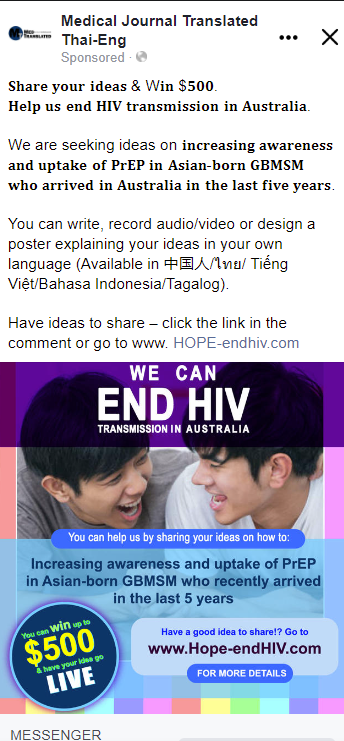

**Figure S1. The Facebook advertisement for the open call in Australia**

**Table S3. The score sheets for the submissions from the open call in Australia**

| **Rank** | **Idea** | **Total** |
| --- | --- | --- |
| 1 | Anonymous, online service where Asian-born GBMSM can both educated themselves on PrEP and it's applications, while at the same time being a safe portal for obtaining PrEP for themselves. | 26.8 |
| 2 | Media campaigns about PrEP and STI clinics in Vietnamese run in popular Vietnamese social media. | 25.2 |
| 3 | Poster to increase PrEP awareness (with a pic) | 24.8 |
| 4 | Give all the Asian-born homosexual people free access to PREP (a trial so they can see how effective it is) and they can decide if they want to continue using it. | 23 |
| 5 | 1. advertising in places frequented by or popular with newly arrived Asian-born GBMSM 2. Find, coach & advertise bilingual GP's to prescribe PrEP.  3. The Australian government need to make PrEP more accessible for migrant by providing subsidized PrEP | 22.8 |
| 6 | Social Media Idea: short videos on Tiktok/Instagram/Wechat, share their PreP bottle with a designed hashtag, work with University | 22.6 |
| 7 | -Social media platforms (e.g. YouTube and TikTok influencers, WeChat) -Poster/billboard advertising in areas frequented by target groups | 17.8 |
| 8 | Stop just advertising to specific groups of people and advertise to all people. STI's need to be looked at just like any other disease or infection. | 16.8 |
| 9 | implement mandatory 2nd yearly blood test analysis for every citizen of Australia and every overseas arrival upon entry | 10.8 |

**The full details of the top three submissions in the open call in Australia**

- **The first prize**

To increase uptake of PrEP among GBMSM, it is important to first identify potential barriers that GBMSM face in taking up PrEP. Evidence shows that dominant thematic barriers include stigma, inaccessibility of health systems as well as low HIV risk perception, amongst others. Such barriers are highly pertinent to the population of Asian-born GBMSM who arrived in Australia in the last 5 years. Besides unfamiliarity with the Australian healthcare system contributing to upstream barriers that keep health systems inaccessible to this population, a cultural background of conservativism feeds strongly into stigma as well. Furthermore, many individuals from this particular background come from home countries where HIV and/or sexual health is heavily stigmatized As such, it is clear that outside of raising awareness to combat low HIV risk perception amongst this population, a key barrier preventing uptake of PrEP in this population is also stigma. The duality of this issue is supported by the evidence base where a qualitative study of newly arrived Asian-born gay men by Phillips et al. highlighted the dual-pronged issues of stigma and lack of knowledge.
I propose an anonymous, online service where Asian-born GBMSM can both educated themselves on PrEP and it's applications, while at the same time being a safe portal for obtaining PrEP for themselves.

In terms of addressing the issue of a poor knowledge background towards PrEP and perhaps Australian sexual healthcare / healthcare in general, the online service would provide information and education on this topic. In order to increase uptake, the online service should be accessible both on smartphones, laptops and other devices. By addressing inadequacies in the knowledge base of the population, the online service serves to tackle more upstream barriers preventing uptake of PrEP in this population.

Crucially, it is also important to tackle downstream barriers in uptake of PrEP. Asian-born GBMSM face a myriad of factors such as stigmatization of HIV, poor experiences with healthcare in Australia and/or their home country, the need for individuals to maintain privacy and/or shame stemming from background, amongst many others. Besides long-term challenging of stigmatization towards sexual health, it is important to offer Asian-born GBMSM with a solution to uptake PrEP that preserves privacy, maintains dignity and reduces any potential sources of stigma. As such, an online service where GBMSM can relatively anonymously and safely obtain PrEP. The online service would involve a quick consultation with a physician online who would counsel the individual on side effects, other STIs and provide any medical advice required. PrEP can then be prescribed to the patient and discretion can also be practiced through innovative ways such as using one-time links to send prescriptions which ensures patients donâ€™t have to give out their personal emails, if they do not wish to do so. In dealing with multiple cultural backgrounds, it is also pertinent that the physician on the other side of the screen be trained in culturally safe healthcare consulting and hence provide better outcomes for Asian-born GBMSM arriving in Australia within the last 5 years.

- **The second prize**

Initiatives to improve Vietnamese speaking gay access to PREP

Context: There are gaps between: the reduction in new HIV notifications was smaller for the Australian born men who have ses with men (MSM)  13% versus 33% of overseas-born peers (Aung et al, 2020); higher percentage of later stage in HIV diagnosis and number of people who never had STI test previously in overseas born groups (Aung et al, 2020); increasing proportion of undiagnosed HIV (Patel et al, 2021); and the majority of HIV notifications in 2018 were from men born in Asia (54%, up from 32% in 2009) (Philpot et al, 2021). It is worth considering some initiatives to improve these inequality in overseas-born MSM access to healthcare in general and PREP specifically

Aim: this initiative aims to reach Vietnamese-born MSM, including international students Vietnamese MSM, newly arrival MSM in Sydney. Advocate for their needs to improve access to sexual health care; Increase their HIV testing and; Improve their engagement with local health care services; provide information/knowledge and facilitate their access to PREP/PEP;

Possible perceived barriers: Low risk perceptions about STI/HIV; Cost of STI/HIV testing. Unaware of PREP or perceived barriers to access PREP like PREP purchasing cost as not Medicare card holders, side effects of PREP; Language barriers; Privacy/confidentiality; Fear of stigma/discrimination; Not aware of the services and/or how to navigate Australian healthcare system

Initiatives: Media campaigns about PREP and STI clinics in Vietnamese run in popular Vietnamese social media. Given Facebook the most popular social network in Vietnam and its easily facilitating of any internationally promotional programs  versus other domestic popular media which does not support overseas activities, most popular Facebook  communities for Vietnamese in Australia,  such as Vietnamese in Sydney - 30K members; Dinh Cu & Cuoc Song Uc (Immigration and Life in Australia) - 63K members, Vietnamese Students in Australia - 73K members; Vietnamese Australian Student Association - 70K members, VASA - Vietnamese Australian student Association - Melbourne 86K,  Vietnamese Students In Sydney - 72K, SIA - Vietnamese Students In Australia 38K, etc should be considered.  These groups are the main sources for any Vietnamese new migrants to exchange information and learn about new lives in Australia. With Vietnamese traditional cultural strait is still dominant, audience targeting Facebook ads would give more reassurance re confidentiality and privacy. Other main features of STI services such as confidential/privacy, free of charge, Vietnamese interpreters, etc should be emphasized in the content of the campaigns. Word by mouth is also a way for Vietnamese people to spread information. By that I meant the number of people knowing about PREP not limited to the members of these Facebook groups

- **The third prize**

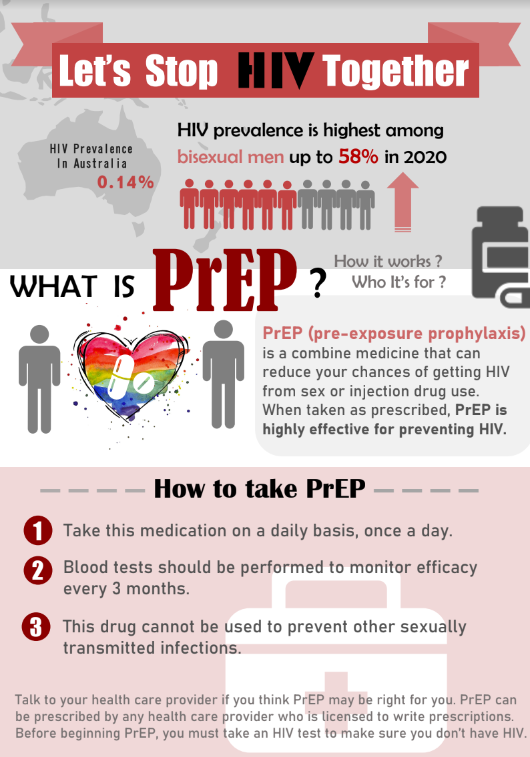

**Supplementary 3. Additional details on the open call in the Philippines**

The open call submission was open from October 5 to November 2, 2022. To ensure the smooth execution of the crowdsourcing call, the project team established two committees: the organizing committee and the steering committee. The six-member steering committee oversaw the overall process and ensured that the call adhered to the predetermined criteria. They also served as the final selection judges who evaluated the entries and determined the winners. Whereas the project team served as the organizing committee, overseeing the day-to-day operations of the crowdsourcing call. They were responsible for promoting the call, receiving and managing submissions, and preparing the entries for judging. The steering (six members) and organizing (eight project team members, four consultants) committees comprised clinicians, academics, researchers, social scientists, community-based service providers, and representatives from the LGBTQIA community.

The project team utilized various communication and promotion strategies (including having its own social media page <https://www.facebook.com/EndHIV.HOPEPh>) to encourage broad participation and diverse submissions. The team leveraged its community networks and affiliations and formed strategic partnerships with key government agencies to disseminate the call through social media. A social media plan was also created and shared with partners to streamline all shared materials and posts in the table below. The promotional materials for the call included the poster and video for the crowdsourcing call, countdown posts, and weekly social media cards. Some social media cards were boosted to broaden the call's reach further and attract quality submissions.

The project team accepted entries through email, online form, and in-person or courier submission to ensure accessibility. Submitted entries underwent a two-step screening process, starting with an initial eligibility screening by the project team. Screening of entries was done to ensure that the eligibility criteria (i.e., developed by Filipinos or lead/core members being Filipinos, focusing on increasing PrEP uptake among key populations, and submitting a complete entry form within the set deadline) were all met. Eligible entries then underwent a final scoring and evaluation by the expert panel of judges.

The expert panel of judges, composed of the steering committee and three additional experts, scored the entries based on predetermined criteria, including appropriateness, innovativeness, inclusiveness, affordability, potential effectiveness, potential scalability, and potential sustainability. Each entry was randomly assigned to two judges for final scoring and evaluation. The judges' scores were averaged, and the weighted average scores were sorted from highest to lowest to determine the top ten finalists. The top three entries were then recognized as the winners. The first, second, and third placers received a plaque of recognition and a cash incentive of Php 80,000 (AUD$ 2,300); Php 40,000 (AUD$ 1,160); and Php 20,000 (AUD$ 580), respectively. The winners and finalists were contacted via email to coordinate the awarding of their prizes.

The winners and finalists were publicly announced on the project’s social media pages on December 6, 2022.^44^ The project acknowledged the names of the innovators/submitters, or the name/s of projects or ideas submitted, depending on their preference. The social media post was also shared with the project partner sites for further dissemination. The project team celebrated the top three winners through a formal awards ceremony during the hackathon event (Hack for PrEP) in January 2023.^42^ They were also given an opportunity to present their proposed solution during the hackathon in front of the experts and the three partner sites where we intend to pilot-test an innovative solution that can potentially increase PrEP use.

**Table S4. The purposed social media advertisement plan for the open call in the Philippines**

| **HOPE Philippines Crowdsourcing Call 2022**  **Social Media Content Plan** | | | | |
| --- | --- | --- | --- | --- |
| **Week 1** | | | | |
| **Date and Time** | **Social Media Card/**  **Promotional Material Link** | **Facebook Caption** | **Twitter Caption** | **Notes** |
| **October 4** | [Teaser Video](https://drive.google.com/file/d/1IZ9EHLIH4ZSZSdgsOAeeGgmHHEyYP0cu/view?usp=sharing) | Something exciting is coming tomorrow!   Like and follow our accounts to stay updated!  **Facebook Page:** <https://www.facebook.com/EndHIV.HOPEPh>  **Twitter:** @EndHIV_HOPEPh | Something exciting is coming tomorrow!   Like and follow our accounts to stay updated! **Facebook Page:** <https://www.facebook.com/EndHIV.HOPEPh>  **Twitter:** @EndHIV_HOPEPh | - Posted |
| **October 5  *1 PM*** | [Poster](https://drive.google.com/file/d/19OorGBViFxiA3w3rH-_wsaVXGJc30rp1/view?usp=sharing) | Call for Submissions: The search is on for solutions that increase PrEP use to help end HIV transmission in the Philippines!  If you have an idea or an innovative solution, we want to hear from you! Submit your entry on or before October 19, 2022 at 11:59 PM.  **Scan QR Code or click this link:**  [**https://bit.ly/HOPEPhOnlineSubmissionForm**](https://bit.ly/HOPEPhOnlineSubmissionForm?fbclid=IwAR3IKxB4FZedWdOiJYpRAUMvqC-bAwb9U4_zESV2chEL-fLcxontozcS7u8)  **Or download form here and submit as PDF file:**  [**https://bit.ly/HOPEPhCallSubmissionForm**](https://bit.ly/HOPEPhCallSubmissionForm)  **For more details, visit:** <https://bit.ly/HOPEPhOpenCallFAQs>  **Got more questions? Message us through:**  **Facebook:** <https://www.facebook.com/EndHIV.HOPEPh>  **Twitter:** https://twitter.com/EndHIV_HOPEPh  **Email**:  #PrEPTogether #SpreadHOPE | The search is on for solutions that increase PrEP use to help end HIV transmission in the Philippines!  If you have an idea or an innovative solution, we want to hear from you! Scan QR to submit your entry on/before October 19, 2022 (11:59 PM)  #PrEPTogether #SpreadHOPE | - Posted - Updated 10/13/2022 |
| **Week 2** | | | | |
| **Date** | **Social Media Card/**  **Promotional Material Link** | **Facebook Caption** | **Twitter Caption** | **Notes** |
| **October 11**  ***12 PM*** | [Final Crowdsourcing Video](https://drive.google.com/file/d/1FbdSHAJzdcPL68fyCdGzdHBnAHS-bHqR/view?usp=sharing) | The search is on for solutions that increase PrEP use to help end HIV transmission in the Philippines!  If you have a new idea, working prototype, or a small to medium scale implementation for six months or more aimed at increasing PrEP uptake among key populations – we want to hear from you!  Submit your entry on or before October 19, 2022 at 11:59 PM.  **Scan QR Code or click this link:**  [**https://bit.ly/HOPEPhOnlineSubmissionForm**](https://bit.ly/HOPEPhOnlineSubmissionForm?fbclid=IwAR3IKxB4FZedWdOiJYpRAUMvqC-bAwb9U4_zESV2chEL-fLcxontozcS7u8)  **Or download form here and submit as PDF file:**  [**https://bit.ly/HOPEPhCallSubmissionForm**](https://bit.ly/HOPEPhCallSubmissionForm)  **For more details, visit:** <https://bit.ly/HOPEPhOpenCallFAQs>  **Got more questions? Message us through:**  **Facebook:** <https://www.facebook.com/EndHIV.HOPEPh>  **Twitter:** https://twitter.com/EndHIV_HOPEPh  **Email**:  #PrEPTogether #SpreadHOPE | Call for submission!   If you have a new idea, working prototype, or a small to medium scale implementation aimed at increasing PrEP uptake – we want to hear from you!  Submission Deadline: Oct. 19, 2022 (11:59 PM)  #PrEPTogether #SpreadHOPE | - Video Link updated (10/10/22) - Posted |
| **October 12**  ***1 PM*** | [What is PrEP?](https://drive.google.com/file/d/1fuHglPxTr4dnA08x-noEX0_zdHlHE2kg/view?usp=sharing) | Call for submissions!  The search is on for solutions to increase PrEP use towards ending HIV transmission in the Philippines. But wait, what is PrEP?    **Scan QR Code or click this link:**  [**https://bit.ly/HOPEPhOnlineSubmissionForm**](https://bit.ly/HOPEPhOnlineSubmissionForm?fbclid=IwAR3IKxB4FZedWdOiJYpRAUMvqC-bAwb9U4_zESV2chEL-fLcxontozcS7u8)  **Or download form here and submit as PDF file:**  [**https://bit.ly/HOPEPhCallSubmissionForm**](https://bit.ly/HOPEPhCallSubmissionForm)  **For more details, visit:** <https://bit.ly/HOPEPhOpenCallFAQs>  **Got more questions? Message us through:**  **Facebook:** <https://www.facebook.com/EndHIV.HOPEPh>  **Twitter:** https://twitter.com/EndHIV_HOPEPh  **Email**:  #PrEPTogether #SpreadHOPE | Call for submissions!  The search is on for solutions to increase PrEP use to end HIV transmission in the Philippines. But wait, what is PrEP?  For details, visit: <https://bit.ly/HOPEPhOpenCallFAQs>  Got questions?   #PrEPTogether #SpreadHOPE | - Posted |
| **October 13**  **1 PM** | [Updated Poster](https://drive.google.com/file/d/1Sui08W3XUMgFaSsdirStdH70Fe-g1FzS/view?usp=sharing) | Call for Submissions: The search is on for solutions that increase PrEP use to help end HIV transmission in the Philippines!  If you have an idea or an innovative solution, we want to hear from you! Submit your entry on or before October 19, 2022 at 11:59 PM.  **Scan QR Code or click this link:**  [**https://bit.ly/HOPEPhOnlineSubmissionForm**](https://bit.ly/HOPEPhOnlineSubmissionForm?fbclid=IwAR3IKxB4FZedWdOiJYpRAUMvqC-bAwb9U4_zESV2chEL-fLcxontozcS7u8)  **Or download form here and submit as PDF file:**  [**https://bit.ly/HOPEPhCallSubmissionForm**](https://bit.ly/HOPEPhCallSubmissionForm)  **For more details, visit:** <https://bit.ly/HOPEPhOpenCallFAQs>  **Got more questions? Message us through:**  **Facebook:** <https://www.facebook.com/EndHIV.HOPEPh>  **Twitter:** https://twitter.com/EndHIV_HOPEPh  **Email**:  #PrEPTogether #SpreadHOPE | Call for Submissions: The search is on for solutions that increase PrEP use to help end HIV transmission in the Philippines!  If you have an idea or an innovative solution, we want to hear from you! Scan QR to submit your entry on/before October 19, 2022 (11:59 PM)  #PrEPTogether #SpreadHOPE | - New Post Added (10/13/2022) - Posted |
| **October 14**  ***1 PM*** | [What is a Crowdsourcing Call?](https://drive.google.com/file/d/1KL4EoUuOqhB5CH3s4KdTNRoFlZapk59W/view?usp=sharing) | Through a crowdsourcing call, we are looking for solutions that increase PrEP use to help end HIV transmission in the Philippines.  Submit your entry on or before October 19, 2022 at 11:59 PM  **Scan QR Code or click this link:**  [**https://bit.ly/HOPEPhOnlineSubmissionForm**](https://bit.ly/HOPEPhOnlineSubmissionForm?fbclid=IwAR3IKxB4FZedWdOiJYpRAUMvqC-bAwb9U4_zESV2chEL-fLcxontozcS7u8)  **Or download form here and submit as PDF file:**  [**https://bit.ly/HOPEPhCallSubmissionForm**](https://bit.ly/HOPEPhCallSubmissionForm)  **For more details, visit:** <https://bit.ly/HOPEPhOpenCallFAQs>  **Got more questions? Message through:**  **Facebook:** <https://www.facebook.com/EndHIV.HOPEPh>  **Twitter:** https://twitter.com/EndHIV_HOPEPh  **Email**:  #PrEPTogether #SpreadHOPE | Through a crowdsourcing call, we are looking for solutions that increase PrEP use to help end HIV transmission in the Philippines.  Want to know more?  **Visit** <https://bit.ly/HOPEPhOpenCallFAQs>    **Got questions?**  Email us at  #PrEPTogether #SpreadHOPE | - SocMed Card Link updated (10/10/22) - Repost on Oct. 14, 2022 (initial Oct 7) |
| **Week 3** | | | | |
| **Date** | **Social Media Card/**  **Promotional Material Link** | **Facebook Caption** | **Twitter Caption** | **Notes** |
| **October 17**  ***1 PM*** | [What solutions can I submit?](https://drive.google.com/file/d/1MjFLbFRN-vtt9ZnHhB55SeDJa3GkYlR0/view?usp=sharing) | Call for submissions!   Wondering what you can submit?  A wide range of solutions regardless of innovation stage, aimed at increasing the use of PreP among key populations, are accepted.  **Scan QR Code or click this link:**  [**https://bit.ly/HOPEPhOnlineSubmissionForm**](https://bit.ly/HOPEPhOnlineSubmissionForm?fbclid=IwAR3IKxB4FZedWdOiJYpRAUMvqC-bAwb9U4_zESV2chEL-fLcxontozcS7u8)  **Or download form here and submit as PDF file:**  [**https://bit.ly/HOPEPhCallSubmissionForm**](https://bit.ly/HOPEPhCallSubmissionForm)  **For more details, visit:** <https://bit.ly/HOPEPhOpenCallFAQs>  **Got more questions? Message us through:**  **Facebook Page:** <https://www.facebook.com/EndHIV.HOPEPh>  **Twitter:** https://twitter.com/EndHIV_HOPEPh  **Email**:  #PrEPTogether #SpreadHOPE | Call for submissions!   Wondering what you can submit? A wide range of solutions regardless of innovation stage, aimed at increasing the use of PreP among key populations, are accepted.  **Got more questions? Email at**  #PrEPTogether #SpreadHOPE | - Posted |
| **October 19**  ***1 PM*** | [Extended Deadline Announcement (Prizes)](https://drive.google.com/file/d/15vS1dMJNKBMM6vE9ZO6sKHsC4_6fFKZq/view?usp=sharing) | We heard you! The deadline for submission is extended until November 2, 2022 (11:59 PM)  Top winners and finalists will receive a cash prize, plaque of recognition, certificate, and an opportunity to participate in the hackathon.  **Scan QR Code or click this link:**  [**https://bit.ly/HOPEPhOnlineSubmissionForm**](https://bit.ly/HOPEPhOnlineSubmissionForm?fbclid=IwAR3IKxB4FZedWdOiJYpRAUMvqC-bAwb9U4_zESV2chEL-fLcxontozcS7u8)  **Or download form here and submit as PDF file:**  [**https://bit.ly/HOPEPhCallSubmissionForm**](https://bit.ly/HOPEPhCallSubmissionForm)  **For more details, visit:** <https://bit.ly/HOPEPhOpenCallFAQs>  **Got more questions? Message us through:**  **Facebook:** <https://www.facebook.com/EndHIV.HOPEPh>  **Twitter:** https://twitter.com/EndHIV_HOPEPh  **Email**:  #PrEPTogether #SpreadHOPE | We heard you! The deadline for submission is extended until Nov. 2, 2022 (11:59 PM)  Top winners & finalists will receive a cash prize, plaque/certificate of recognitions, and an opportunity to participate in a hackathon.  Scan QR to submit now!  #PrEPTogether #SpreadHOPE | - SocMed Card Link updated (10/13/22) - ~~Posted~~ |
| **Oct 19**  **5PM** | [Poster (Extended Deadline Version)](https://drive.google.com/file/d/1xp5mqyhtDozdLOOtNM8aQZZeTBBn1miT/view?usp=sharing) | 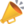 The deadline for submission is extended until November 2, 2022 (11:59 PM)   Don’t miss this chance! The search is still on for solutions that increase PrEP use to help end HIV transmission in the Philippines!  If you have an idea or an innovative solution, we want to hear from you!   **Scan QR Code or click this link:**  [**https://bit.ly/HOPEPhOnlineSubmissionForm**](https://bit.ly/HOPEPhOnlineSubmissionForm?fbclid=IwAR3IKxB4FZedWdOiJYpRAUMvqC-bAwb9U4_zESV2chEL-fLcxontozcS7u8)  **Or download form here and submit as PDF file:**  [**https://bit.ly/HOPEPhCallSubmissionForm**](https://bit.ly/HOPEPhCallSubmissionForm)  **For more details, visit:** <https://bit.ly/HOPEPhOpenCallFAQs>  **Got more questions? Message us through:**  **Facebook:** <https://www.facebook.com/EndHIV.HOPEPh>  **Twitter:** https://twitter.com/EndHIV_HOPEPh  **Email**:  #PrEPTogether #SpreadHOPE | 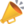Extended Deadline: November 2, 2022 (11:59 PM)  Don’t miss this chance! The search is still on for solutions that increase PrEP use to help end HIV transmission in the Philippines!  If you have an idea or an innovative solution, we want to hear from you! Scan QR to submit your entry.  #PrEPTogether #SpreadHOPE | - Posted |
| **Week 4** | | | | |
| **Date** | **Social Media Card/**  **Promotional Material Link** | **Facebook Caption** | **Twitter Caption** | **Notes** |
| **October 26**  ***1 PM*** | [7 days to go (How to Submit?)](https://drive.google.com/file/d/1B-QkKCX1rdnMBQ29iVmDLqkqtaXY1rbg/view?usp=sharing) | SEVEN days to go! Send in your submissions now!  Extended Deadline: November 2, 2022 (11:59 PM)  **Scan QR Code or click this link:**  [**https://bit.ly/HOPEPhOnlineSubmissionForm**](https://bit.ly/HOPEPhOnlineSubmissionForm?fbclid=IwAR3IKxB4FZedWdOiJYpRAUMvqC-bAwb9U4_zESV2chEL-fLcxontozcS7u8)  **Or download form here and submit as PDF file:**  [**https://bit.ly/HOPEPhCallSubmissionForm**](https://bit.ly/HOPEPhCallSubmissionForm)  **For more details, visit:** <https://bit.ly/HOPEPhOpenCallFAQs>  **Got more questions? Message us through:**  **Facebook:** <https://www.facebook.com/EndHIV.HOPEPh>  **Twitter:** https://twitter.com/EndHIV_HOPEPh  **Email**:  #PrEPTogether #SpreadHOPE | 7 days to go! Send in your submissions now!  Extended Deadline: November 2, 2022 (11:59 PM)  **For more details, visit:** <https://bit.ly/HOPEPhOpenCallFAQs>  **Got more questions? Email us at**  #PrEPTogether #SpreadHOPE | - Posted |
| **October 28  *1 PM*** | [5 days to go (Why Submit?)](https://drive.google.com/file/d/14GzXPaXt6RZRP2Q03esBznQ4WZiJu_Sf/view?usp=sharing) | FIVE days to go! Take part in addressing the growing problem of HIV transmission in the country. Send your entries now!  Extended Deadline: November 2, 2022 (11:59 PM)  **Scan QR Code or click this link:**  [**https://bit.ly/HOPEPhOnlineSubmissionForm**](https://bit.ly/HOPEPhOnlineSubmissionForm?fbclid=IwAR3IKxB4FZedWdOiJYpRAUMvqC-bAwb9U4_zESV2chEL-fLcxontozcS7u8)  **Or download form here and submit as PDF file:**  [**https://bit.ly/HOPEPhCallSubmissionForm**](https://bit.ly/HOPEPhCallSubmissionForm)  **For more details, visit:** <https://bit.ly/HOPEPhOpenCallFAQs>  **Got more questions? Message us through:**  **Facebook:** <https://www.facebook.com/EndHIV.HOPEPh>  **Twitter:** https://twitter.com/EndHIV_HOPEPh  **Email**:  #PrEPTogether #SpreadHOPE | FIVE days to go! Take part in addressing the growing problem of HIV transmission in the country. Send your entries now!  Extended Deadline: November 2, 2022 (11:59 PM)  Got more questions? Email us at  #PrEPTogether #SpreadHOPE | - SocMed Card Link updated (10/10/22) - SocMed Card Link updated (10/13/22) - Posted |
| **Week 5** | | | | |
| **Date** | **Social Media Card/**  **Promotional Material Link** | **Facebook Caption** | **Twitter Caption** | **Notes** |
| **November 1  *1 PM*** | [1 day to go (Who can join?)](https://drive.google.com/file/d/1rbyUCEigM8PYtRxGrb_bEM1K8ez6T-R8/view?usp=sharing) | ONE day to go! We are still accepting submissions to the call for solutions to increase PrEP use in the Philippines!  This call is open to all individuals, groups, and organizations in the Philippines. Send in your submissions now!  Extended Deadline: November 2, 2022 (11:59 PM)  **Scan QR Code or click this link:**  [**https://bit.ly/HOPEPhOnlineSubmissionForm**](https://bit.ly/HOPEPhOnlineSubmissionForm?fbclid=IwAR3IKxB4FZedWdOiJYpRAUMvqC-bAwb9U4_zESV2chEL-fLcxontozcS7u8)  **Or download form here and submit as PDF file:**  [**https://bit.ly/HOPEPhCallSubmissionForm**](https://bit.ly/HOPEPhCallSubmissionForm)  **For more details, visit:** <https://bit.ly/HOPEPhOpenCallFAQs>  **Got more questions? Message us through:**  **Facebook:** <https://www.facebook.com/EndHIV.HOPEPh>  **Twitter:** https://twitter.com/EndHIV_HOPEPh  **Email**:  #PrEPTogether #SpreadHOPE | Call for submissions: ONE day to go!  This call is open to all individuals, groups, and organizations in the Philippines. Hurry, send in your submissions now!  Got more questions? Email us at  #PrEPTogether #SpreadHOPE | - SocMed Card Link updated (10/13/22) - Posted |
| **November 2  *8 AM*** | [Last day of submission](https://drive.google.com/file/d/1-aSRwowmiatzlFv4AntVM9DXjRmbgpUN/view?usp=sharing) | Don’t miss this chance! Today is the LAST DAY to submit your ideas and innovative solutions!  Submit your entries now!  **Scan QR Code or click this link:**  [**https://bit.ly/HOPEPhOnlineSubmissionForm**](https://bit.ly/HOPEPhOnlineSubmissionForm?fbclid=IwAR3IKxB4FZedWdOiJYpRAUMvqC-bAwb9U4_zESV2chEL-fLcxontozcS7u8)  **Or download form here and submit as PDF file:**  [**https://bit.ly/HOPEPhCallSubmissionForm**](https://bit.ly/HOPEPhCallSubmissionForm)  **For more details, visit:** <https://bit.ly/HOPEPhOpenCallFAQs>  **Got more questions? Message us through:**  **Facebook:** <https://www.facebook.com/EndHIV.HOPEPh>  **Twitter:** https://twitter.com/EndHIV_HOPEPh  **Email**:  #PrEPTogether #SpreadHOPE | Don’t miss this chance! Today is the LAST DAY to submit your ideas and innovative solutions!  Scan QR code to submit now!  Got more questions? Email us at  #PrEPTogether #SpreadHOPE | - SocMed Card Link updated (10/13/22) - Posted |
| **November 2  11:59 PM** | [End of Submission](https://drive.google.com/file/d/1EEya68VqQW6TpXU86zEPGkXoM1JsdbCo/view?usp=sharing) | Submission of entries is now closed! Submissions will now be evaluated by a panel of judges. Stay tuned for the announcement of winners and finalists.  Like and follow us on:  **Facebook:** <https://www.facebook.com/EndHIV.HOPEPh>  **Twitter:** https://twitter.com/EndHIV_HOPEPh  #PrEPTogether #SpreadHOPE | Submission of entries is now closed! Submissions will now be evaluated by a panel of judges. Stay tuned for the announcement of winners and finalists.  Like and follow us on:  **Facebook:** <https://www.facebook.com/EndHIV.HOPEPh>  **Twitter:** www.twitter.com/EndHIV_HOPEPh  #PrEPTogether #SpreadHOPE | - SocMed Card Link updated (10/13/22) |

**Table S5. The full details of the top three submissions in the open call in the Philippines**

| RANK | PROJECT TITLE | **Innovator** | **Brief Course Description** |
| --- | --- | --- | --- |
| 1 | [**Project SHAFT**](https://docs.google.com/document/d/1igXZnzGXramSTmEXQ8WqFBuyKkhrlRH3/edit?usp=share_link&ouid=114056967177128116390&rtpof=true&sd=true) | **Gerald** | Project SHAFT - Sustainable HIV & AIDS Awareness through Fashion Tales aims to partner with a design school to create sustainable graphical, digitally printed ready-to-wear garments that would include the artistry of artists and fashion design students. These designs should evoke curiosity in everyone, to inform on what PREP is, what it does, and to further seek why it is important in today's society. |
| 2 | [**Community-Based Mobilization Algorithm**](https://docs.google.com/document/d/17QL5BbX-bT1LKHhZHx_JwEbWDNHwK-4K/edit?usp=share_link&ouid=114056967177128116390&rtpof=true&sd=true) | Niel Gabriel Saplagio | This innovation utilizes a recommender algorithm similar to video streaming platforms that recommend certain shows to viewers. This algorithm optimizes the mobilization events for key populations based on the data of current community-based services such as HIV Testing and PrEP and other publicly available data. |
| 3 | [**PharmAssist for PrEP**](https://docs.google.com/document/d/1MDgfuOZdonboPBcTQqj7xza9KL0qBuqt/edit?usp=share_link&ouid=114056967177128116390&rtpof=true&sd=true) | Charles Mandy G. Ayran | PharmAssist for PrEP is a three-phased proposed solution aimed at increasing the recognition of the clinical benefits of PrEP and normalizing its use through the (1) development of a universal PrEP access QR code and symbol, which will be linked to the (2) development of a mobile application containing medication information about PrEP use, with instructions on the nearest PrEP access and dispensing points, and (3) expansion of access points through enrolment of patients to the community pharmacy of their choice. |
| 4 | [**Holistic Strategy for PrEP Promotion and Adherence**](https://docs.google.com/document/d/1DBvK_72FW7d1GEcHL2Xsx0JY9eZQDqGx/edit?usp=share_link&ouid=114056967177128116390&rtpof=true&sd=true) | TEAM OF  DONNA C. ASAN | A solution that modifies existing strategies and introduces new strategies to boost PrEP awareness and adherence throughout the PrEP journey. The holistic strategy focuses on awareness and promotions, consultation and testing, counselling and dispensing, PrEP intake, regular visits and ending medication. |
| 5 | [**The Untapped Pharmacist in HIV Prevention Program**](https://drive.google.com/file/d/1pA9XPjxrFrZ3uC2lXKYs4v1_yYOR5z7H/view?usp=share_link) | TEAM of Kenneth Aguilar, RN, MAN (Lead Member),  Marilou L. Pascual, RPh,  MPM, PhD., Jem Marie L. Magno, RPh, ClinPharm. | The Untapped Pharmacist HIV Prevention Program comprises a series of phases that focuses on taking advantage of the accessibility, underutilization, and empowerment of community pharmacies and pharmacists to help improve awareness in prevention treatment and stigma reduction. |
| 6 | [**PrEP-Up**](https://docs.google.com/document/d/1Md2jl4Xc1FnkqQXB5BYfNTKPJ5hbYXS1/edit?usp=share_link&ouid=114056967177128116390&rtpof=true&sd=true) | **Benjie M. Clemente** | PrEP-Up aims to develop a mobile application that provides information on basic knowledge about PrEP and how to access this medicine. This mobile application is a comprehensive HIV prevention tool with basic information and links to resources that involve a combination prevention approach, such as HIV/sexually transmitted infection testing, PrEP information and provider locators, and condom promotion. |
| 7 | [**MASK FOR AWARENESS**](https://drive.google.com/file/d/1rUrsh5-nJInEk_sErqqTNEQ3GFU_wj5n/view?usp=share_link) | Angel Astronomo | Mask for Awareness proposes to utilize a movable mask dispenser that offers free face masks that includes a PrEP infographic inside the packaging, which can be strategically placed in a high-demand establishment, community, or any desired setting to promote PrEP. |
| 8 | [**PrEPR:  PreP Reinforced**](https://docs.google.com/document/d/1RsaSkJMq4zWpalNlC4oN6uzhajGft7GI/edit?usp=sharing&ouid=114056967177128116390&rtpof=true&sd=true) | **Matthew David** | PrEPr is a closed-loop data ecosystem that aims to enable clinics and clients to monitor PrEP issuances. It includes the PrEPr Clinics App to be used in administering delivery of PrEP services, a Clients App that will be developed to facilitate virtual consultations, refill requests, and monitor PrEP use of enrolled clients through push notifications reminders, and a Clients module with a social networking feature where users can interact and match with other PrEP users based on their PrEP use patterns. |
| 9 | [**PoSH! (A streamlined dating mobile application)**](https://docs.google.com/document/d/13ggQsj24pos8i1mtViE2qB9x7q0SEKb9/edit?usp=share_link&ouid=114056967177128116390&rtpof=true&sd=true) | **Ryan V. Labana** | "PoSH" is a mobile application that works like a dating app, with a streamlined agenda of educating Filipinos about PrEP. As the "dating" progresses, the key population become knowledgeable about PrEP, and they begin doing "meet-ups" for face-to-face counselling in the social hygiene clinics (or in any available safe space) where they can also get the pills. |
| 10 | [**CBS Motivators for PrEp!**](https://drive.google.com/file/d/1y2lY1r3AwYTwHJHqP8tiWEjrQY-D4Shp/view?usp=share_link) | TEAM of Marie Gabrielle Motril | CBS Motivators for PrEP focuses on training and dispatching CBS motivators trained to help discuss and assist individuals with HIV testing and preventative measures such as PrEP during discussion sessions. |

**Table S6. The score sheets for the submissions from the open call in the Phillipines**

| **Rank** | **Name of Innovation/solution** | **Total Average Score** |
| --- | --- | --- |
| 1 | Project SHAFT | 4.425 |
| 2 | Community-based mobilization algorithm | 4.188 |
| 3 | PharmAssist for PrEP | 4.150 |
| 4 | Holistic Strategy for PrEP Promotion and Adherence | 3.938 |
| 5 | The Untapped Pharmacist in HIV Prevention Program | 3.838 |
| 6 | PrEP-Up | 3.750 |
| 7 | Mask for Awreness | 3.635 |
| 8 | PrEPR: PrEP Reinforced | 3.563 |
| 9 | PoSH! (A streamlined dating mobile application) | 3.515 |
| 10 | CBS Motivators for PrEP! | 3.475 |

**Supplementary 4. Finalists of the PrEP adherence messaging open call in China**

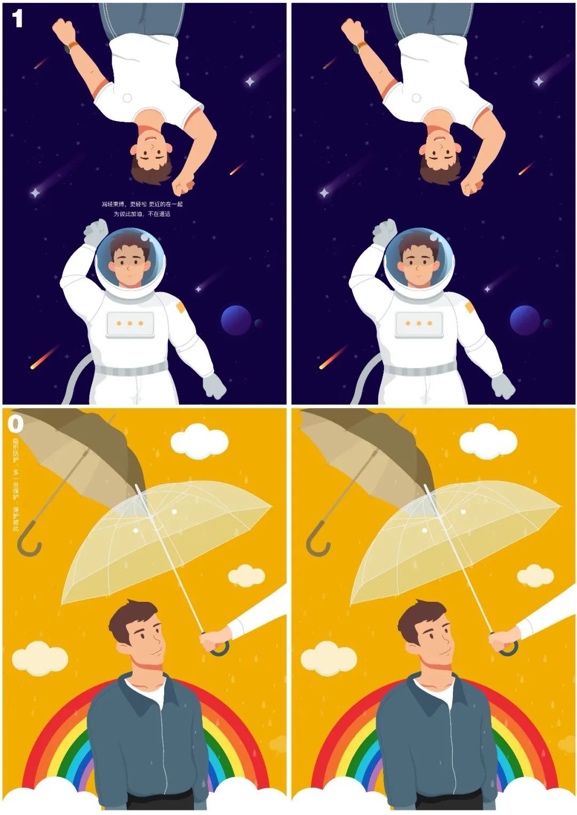

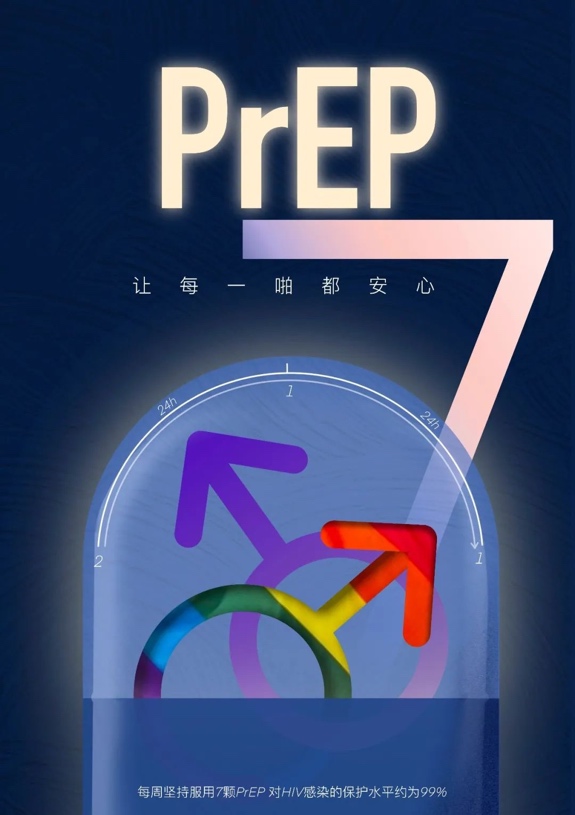

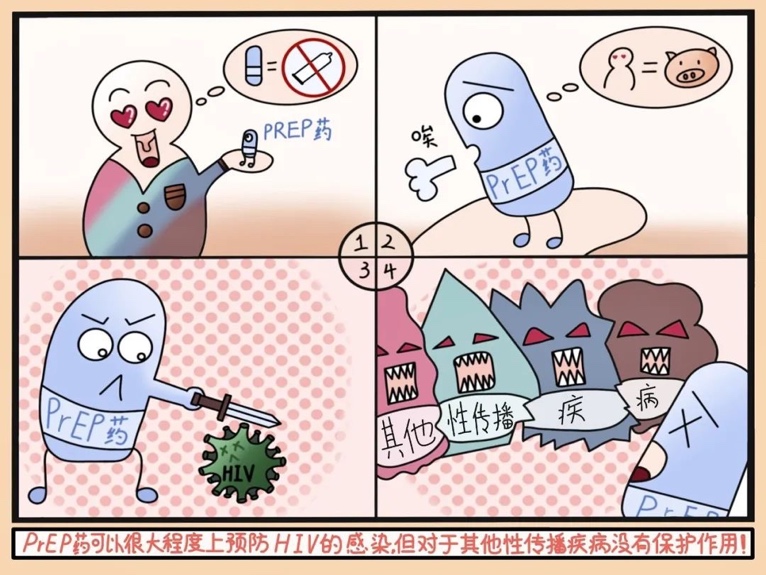

Top left: 1^st^ place

Top right: 2^nd^ place

Bottom left: 3^rd^ place

**Table S7. The scoresheet for the submissions from the open call in China**

| Rank | Idea | Total |
| --- | --- | --- |
| 1 | A poster underscored the effectiveness of PrEP protection. Visual elements include glowing pills, colors, and symbols associated with minority alignment, signifying the community rapport. The time scale and numbers showcase the 2-1-1 dosing strategy and 7-day efficacy of PrEP. | 40.2 |
| 2 | This work emphasizes the effortless and discreet protection offered by PrEP during its usage, aiming to underscore the minimal burden of adopting an additional preventive measure. The visual content also employs positive language to encourage users. | 37.2 |
| * | Three videos were about how to carry medicine, take medicine on time, and regular follow-up. | 37.2 |
| 3 | Comic was used to explain the misunderstandings of PrEP use.  The submission reveals the effect of PrEP on HIV and warns against the misconception that PrEP is a universal STD prevention drug. | 37 |
| 4 | A promotional strip with a pair of hugging hands to wrap the medicine bottle was designed as a clip. It weakened the experience of taking medicine and combined the promotional materials with the medicine bottle so that it was not easy to lose. | 36.8 |
| 5 | The concept of drug packaging is to make the two PrEP regimens clear and straightforward. The design concept can be used not only as promotional picture material on multimedia but also as a reference for the design of innovative PrEP drug packaging. | 34.8 |
| 6 | A PrEP promotional visual material (in the form of a picture) uses an alarm clock, calendar, and post-it notes to make. The aim is to improve the awareness, utilization rate, and medication compliance of PrEP (taking "2+1+1" on demand and daily). | 33.4 |
| 7 | Seven suggestions were put forward mainly for the difficulty of controlling the first dose of event-driven PrEP use, medication-carrying, and habit cultivating. | 31.8 |
| 8 | This work introduces two regimens of PrEP. | 29.6 |
| 9 | The poster was based on a review article published in 2018, and the critical information points of the review were extracted to illustrate the safety and efficacy of PrEP. | 29.2 |
| 10 | Inform people of the importance of PrEP through a brief description. | 28.6 |
| 11 | A single slogan to warn about HIV. | 26.8 |
| 12 | Considering the popularity of shot videos, getting familiar with emoji content and funny short texts is accessible. | 26.4 |
| 13 | Rational use of PrEP to block the virus and lead to a healthy gay life. | 23.2 |
| 14 | Let readers understand the applicable population, safety, administration method, and dependency of PrEP through a picture, and finally leave the two-dimensional code of the article for further understanding. | 22.6 |
| 15 | A single slogan about using PrEP and preventing HIV. | 22.4 |
| 16 | A single slogan to introduce the impact of using PrEP. | 22 |
| 17 | Encourages the applicable population to adhere to the medication and what should be paid attention to adhere to the medication (in the form of a letter) | 21.8 |
| 18 | A single picture without interpretations | 6 |

*: For technical reasons and considering the storage space of the user's phone, this submission was excluded from the final ranking.

**Supplementary 5. The proposed plan on the upcoming open call in Thailand**

In Thailand, the project has not started yet and is waiting for funding from the government and private agencies. We will identify community-driven solutions to optimize PrEP effectiveness and later implement and evaluate solutions through trials, including an economic evaluation. With the current existence of PrEP in the City, a nationwide PrEP campaign implemented by the Institute of HIV Research and Innovation (IHRI) since 2019, the campaign will be delivered under PrEP in the City with a focus in Bangkok and later be scaled up in other prioritized cities.

Before the official launch, preparations will be made simultaneously for the website and social media platforms, alongside the formation of committees consisting of individuals from diverse fields. To ensure the submission of high-quality entries, a series of masterclasses will be organized for prospective participants following the launch. These masterclasses aim to provide additional information, while online interactive sessions will be offered to address any queries they may have. The entry submission period will span two months, during which a comprehensive promotional campaign will be conducted. This campaign will strategically target students from relevant faculties and individuals within networks focusing on health solutions. The submissions can take various formats to showcase solutions related to PrEP effectiveness promotion.

Once the two-month submission period concludes, entry submissions will be closed, and all entries will be showcased on the website, allowing for public votes. The website will feature comment and voting functionalities for one month. The judging panel will subsequently judge the four submissions that receive the most votes to select the top finalist. The judging criteria will include: 1) feasibility, 2) innovation, 3) inclusion, 4) impact and 5) relevancy. The four finalists will be announced and honoured during the award ceremony.

**Table S8. The proposed plan on the upcoming open call in Thailand**

|  | **Thailand (proposed plan)** |
| --- | --- |
| **Problem statement** | Promote the effective use of PrEP among MSM and TGW |
| **Working teams** | Steering and organising committees |
| **Submissions** | Submission form: Online in any formats  Screening: all submissions were included unless not relevant to the topic |
| **Evaluation criteria** | - Feasibility to implement - Innovation of the idea - Provisions of solutions to address the pain points |
| **Judging process** | To be determined |
| **The winner announcement** | IHRI website and social media platforms |
| **Reward** | Cash |
| **Incentives** | To be determined |
